# A Discovery and Verification Approach for Pharmacovigilance using Electronic Health Care Data

**DOI:** 10.1101/2022.05.10.22274885

**Authors:** LJ Dijkstra, T Schink, R Linder, M Schwaninger, I Pigeot, MN Wright, R Foraita

## Abstract

**Introduction:** Pharmacovigilance shifted its focus from spontaneous reporting systems to electronic health care (EHC) data. Usually, a single statistical method is used to detect signals, i.e., potential adverse drug reactions (ADRs).

**Objective and Method:** We present a novel approach to detect ADRs in EHC databases. It aggregates the results of multiple statistical signal detection methods applying Borda count ranking, a preference voting system, which results are used by an expert committee to select plausible signals. The obtained signals are afterwards investigated in tailored pharmacoepidemiological studies to provide support of plausibility or spuriousness of the signal.

We showcase the approach using data from the German Pharmacoepidemiological Research Database on drug reactions of the direct oral anticoagulant rivaroxaban. Results of four statistical methods are aggregated into Borda count rankings: longitudinal Gamma Poisson shrinker, Bayesian confidence propagation neural network, random forests and LASSO. A verification study designed as nested active comparator case-control study was conducted. We included patients diagnosed with atrial fibrillation who initiated anticoagulant treatment with rivaroxaban or with phenprocoumon as active comparator between 2011 and 2017.

**Results:** The case study highlights that our Borda ranking approach (https://borda.bips.eu) is fast, able to retrieve known ADRs and find other interesting signals. Hasty false conclusions are avoided by a verification study, which is, however, time-consuming.

**Conclusion:** Post-market signal detection in EHC data is useful to identify and validate safety signals, particularly a few years after first admission to the market, when spontaneous reports are less frequent and more EHC data are available.

## 1 Introduction

Evidence of the safety of newly approved drugs is often limited. The pivotal randomized clinical trials (RCTs) are powered to assess efficacy so that the sample sizes are too small to examine rare safety outcomes. Patients in RCTs usually have to fulfill several inclusion criteria and especially vulnerable groups such as pregnant women, elderly, and multi-morbid persons are often excluded or underrepresented. Moreover, patients are followed up very closely under controlled conditions over a limited period of time. Therefore, it is essential to also monitor the safety of drugs in routine care.

Traditionally, spontaneous reporting systems form the cornerstone of pharmacovigilance [2-6]. In the last years, research shifted its focus to electronic health care data, which contain drug prescriptions and medical events over time for individual patients [5, 7, 8]. These data sources offer several advantages over spontaneous reporting systems: 1) the total number of patients that were prescribed a drug and/or suffered an adverse drug reaction (ADR) is known, while a spontaneous report is only filed when a drug was prescribed *and* an ADR occurred, 2) under- and over-reporting issues are less pronounced [9, 10], and 3) some electronic health care databases contain follow-up data of several million persons [11].

A plethora of statistical methods and study designs for detecting associations between drugs and ADRs, i.e., signals, in electronic health care databases have been proposed [12-15]. These methods range from disproportionality measures (e.g., reporting odds ratio [16]), hypothesis tests (e.g., Poisson test [17]) and Bayesian shrinkage estimates (e.g., Bayesian confidence propagation neural network, BCPNN [18, 19]) to sparse regression (LASSO [20, 21]). These methods offer different advantages, e.g., disproportionality measures provide risk estimates, sparse regression controls for high-dimensional confounding and the BCPNN can deal with innocent bystanders [13]. The choice of method is non-trivial and depends on many factors [13, 22]. To combine the strengths and overcome the limitations of individual signal detection methods, we propose to employ multiple methods simultaneously and aggregate their individual scores into a single ranking using the Borda count method [23, 24].

After the signal detection phase, the ranking of drug-ADR pairs is usually presented to a committee of medical experts, which triages the signals and decides on any possible actions (e.g., further analyses, issue warnings, request label changes or, in extreme cases, recommend to withdraw the license [25]). We propose to complement signal detection and signal decision with a verification study using state of the art pharmacoepidemiological methods as an additional option to check whether signals persist when standard methods to avoid sources of bias such as controlling for confounding are applied.

We demonstrate the applicability of the approach by means of a case study of the direct oral anticoagulant rivaroxaban (RVX, ATC: B01AF01) using the “German Pharmacoepidemiological Research Database” (GePaRD) [11] which is based on health care claims data.

## 2 Methods

### 2.1 Signal detection of potential adverse drug reactions (ADRs)

Detecting possible adverse drug reactions (ADRs) is commonly done by using a single signal detection method [17-20, 26, 27]. Such methods yield a score for each drug-ADR pair that reflects the ‘strength’ of the association between the drug and the ADR in question [13]. A ranking of the drug-ADR pairs based on these scores is presented to a committee of medical experts for discussion and signal triage. See Figure 1a for a graphical representation of the process.

**Figure 1:**
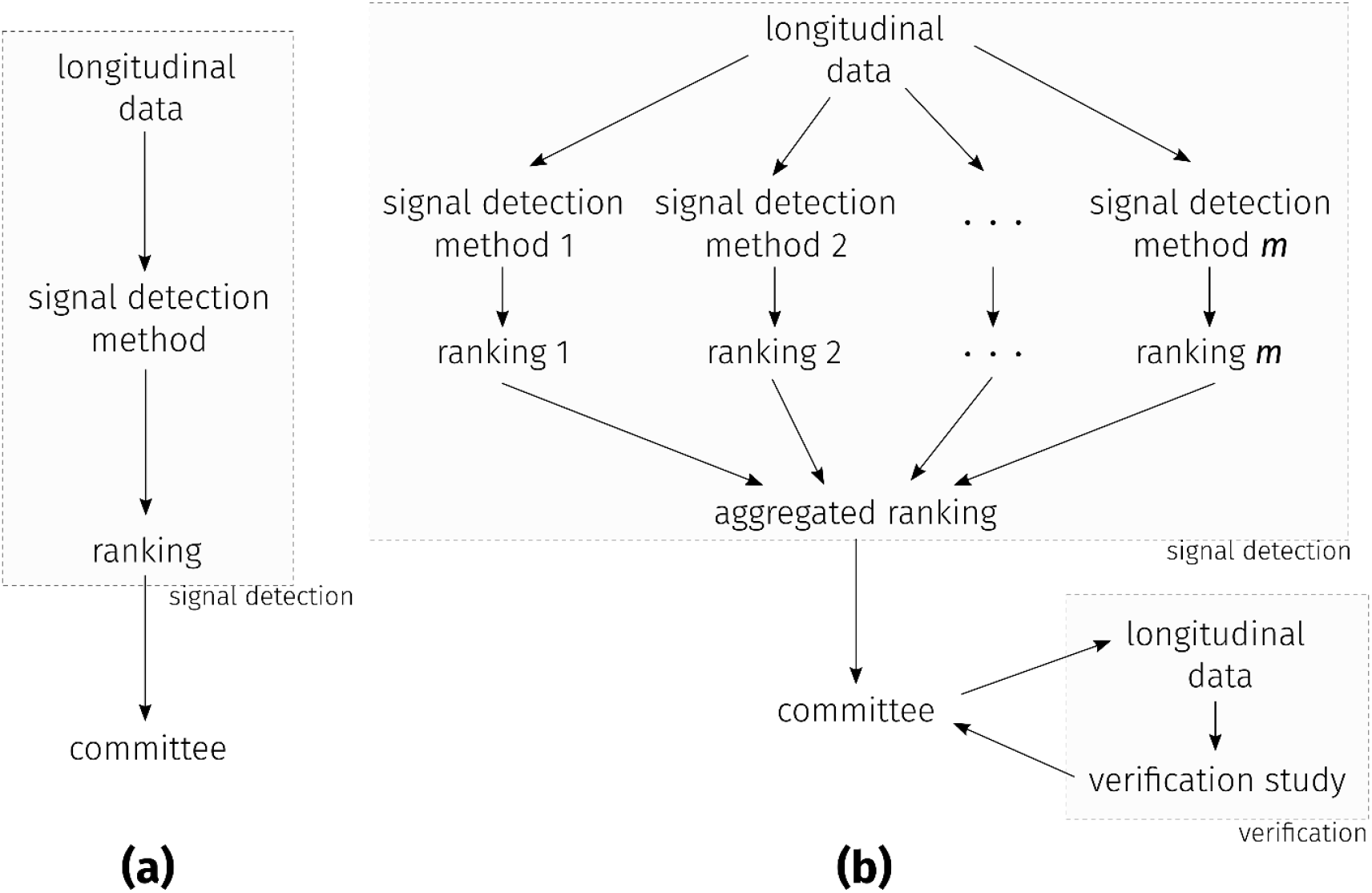
(a) The common approach: signal detection is done by applying a single detection method to the LD, resulting in a single ranking of drugs-ADR pairs. This ranking is presented to a committee of medical experts. (b) The approach presented in this paper: *m* different detection methods are applied to the LD, resulting in *m* rankings. These rankings are aggregated into a single ranking based on the Borda count. The single aggregated ranking is then presented to the committee. In addition, the choice of performing a verification study is available to the committee.

We propose instead to use multiple signal detection methods simultaneously, see Figure 1b. Different methods can yield scores with different interpretations, e.g., the Poisson test [17] yields *p*-values, while the LASSO [20] results in shrunken odds ratio estimates and random forest provides variable importance rankings. Rather than using the raw scores, we aggregate the rankings of the drug-ADR pairs that resulted from the individual methods into a single ranking using the Borda count [23, 24], which is an electoral system for selecting broadly agreed beliefs. The Borda count for each drug-ADR pair is the sum of the ranks for that drug-ADR pair in each of the rankings. The drug-ADR pair with the lowest Borda count is ranked first with rank scored as “1”; the drug-ADR pair with the highest Borda count is placed last. The intuition behind the Borda count ranking is that it captures the ‘consensus’ between the methods rather than the majority. For example, if a drug-ADR pair shows a high level of ‘association’ according to multiple methods, the pair is ranked high in the individual rankings. The sum of the ranks for this pair would be relatively low. As a result, the pair will also be listed high up in the Borda count ranking. Now consider the situation where a pair is ranked high for only a few methods, while the others place it low. This pair would not end up high in the Borda count ranking. The advantage of this approach is that it does not depend on the raw scores directly and can be easily applied, even when the number of methods is large and the assessment criteria of the methods differ. Tournament-style counting is used to account for ties [28].

### 2.2 Signal triage

Let us assume that the Borda count ranking of the drug-ADR pairs is presented to a committee of medical experts. The committee is charged with the task of filtering signals based on the Borda count that might indicate severe and unknown ADRs and taking appropriate actions. Employing expert knowledge in this process is hence paramount. Subject-matter knowledge is required to assess the biological plausibility that an identified ADR is caused by the drug of interest. The possibility of the drug in question to be an innocent bystander, or whether the signal is due to confounding by indication, has to be ruled out. Only considering the top-ranked drug-ADR pairs would be insufficient, since they are likely to contain known ADRs, while unknown ADRs are likely lower on the list. Due to the large number of pairs, it would be infeasible to systematically verify all signals using traditional pharmacoepidemiological studies. Our approach, therefore, is limited to verifying a smaller set of drug-ADR pairs. Instead of individual signals, the committee could also opt to further investigating an outcome in which the diagnostic codes of multiple signals are aggregated into events of interest. There may, however, be medical or pharmacological reasons to expand an event definition from the signal triage to include other diagnostic codes that were not particularly conspicuous in signal detection.

We created an online tool for performing signal triage based on Borda counts, which is available on https://borda.bips.eu. The R-Shiny app computes the Borda count ranks based on uploaded data and helps with graphical representations to select drug-diagnostic code pairs.

### 2.3 Verification of signals

In the third step, the robustness of the filtered signals is tested by an observational study targeted to the specific exposure-outcome relationship of interest. This study should be designed to minimize bias and control for confounding, e.g., use a new user active comparator approach [29], assess all available potential confounders and use adequate statistical methods (see Supplementary Material for more information).

### 2.4 Case study

#### Data Source

For the verification study we used the German Pharmacoepidemiological Research Database (GePaRD). GePaRD is based on claims data from four statutory health insurance (SHI) providers in Germany and currently includes information on approximately 25 million persons who have been insured with one of the participating providers since 2004 or later [11]. Diagnoses are coded based on the 10^th^ version of the International Classification of Diseases in the German Modification (ICD-10-GM). Persons who were insured in the Techniker Krankenkasse between January 2015 and December 2016 were included in the signal detection study. The verification study included patients diagnosed with atrial fibrillation who started anticoagulant treatment with rivaroxaban or with phenprocoumon as the active comparator between 2011 and 2017.

#### Exposure of interest

Rivaroxaban (RVX, ATC: B01AF01) is a direct oral anticoagulant (DOAC) that has been approved for the prevention of stroke and systemic embolism in patients with atrial fibrillation, the treatment of deep vein thrombosis and pulmonary embolism, and the prevention of recurrent deep vein thrombosis and pulmonary embolism in adult patients, and for the prevention of venous thromboembolism in adult patients undergoing elective hip or knee replacement surgery. As is the case with other anticoagulants, clinical studies of rivaroxaban identified hemorrhage as an important safety outcome [30].

GePaRD and the cohort of the signal detection study are described in detail in the supplementary material.

#### Signal detection methods

We employ four signal detection methods: the Bayesian confidence propagation neural network (BCPNN; [19]), the longitudinal Gamma Poisson shrinker (LGPS; [31]), the LASSO [20] and random forests (RF; [32]). Both the BCPNN and the LGPS are univariate methods yielding scores based on 2×2 contingency tables, one for each ADR, see Table 1. The LASSO and the RF are multivariate methods and can take a large number of covariates, e.g., diagnoses and patient information, into account. The scores for the ADRs of the LASSO are defined as the highest penalization parameter for which that ADR enters the active set [33]. The output of the RF is a rank vector of the variable importance score [34].

**Table 1:** A 2×2 table for a drug and ADR.

|  | ADR occurs | ADR does not occur | Total |
| --- | --- | --- | --- |
| Rivaroxaban was prescribed | $a$ | $b$ | $a + b$ |
| Rivaroxaban was not prescribed | $c$ | $d$ | $c + d$ |
| Total | $a + c$ | $b + d$ | $N = a + b + c + d$ |

The count $a$ is the number of time points (in case of GePaRD, quarters) the patients were prescribed rivaroxaban and experienced the ADR in question. The count $b$ is the number of quarters the patients were prescribed the rivaroxaban but did not experience the ADR. The count $c$ and $d$ are constructed in a similar fashion. Note that $N = a + b + c + d$ is the number of the observed quarters of all patients.

A self-controlled case series design [35] was used for the LASSO and the RF, where we considered the first control period, in which the patient was not exposed to the drug, and the first risk period, in which the person was exposed. See Figure S1 in the Supplementary Material for a graphical representation. We noted whether or not the person experienced the ADR in question during the control period and in the risk period. In our analysis, we used the covariates age, sex, and all diagnoses and prescribed medications to account for potential interactions. The ranking of all four methods were than aggregated into Borda count ranking.

#### Signal triage

A committee of pharmacoepidemiologists, pharmacologists, physicians and statisticians reviewed the rankings and discussed plausibility, severity and novelty of the signal.

#### Verification study

The verification study was designed as an active comparator case-control study nested in a cohort of new users of RVX and phenprocoumon (PPC, ATC: B01AA04, the most frequently used vitamin K antagonist in Germany), with atrial fibrillation.

Cases were defined as patients with a diagnosis of the respective outcome of interest. Each case was matched with up to 10 controls by sex, age at index day (± 1 year) and statutory health insurance provider using risk set sampling with time in cohort as the time axis to ensure a similar follow-up as for the corresponding case. Eligible patients hospitalized for any reason at the index date of the case were excluded from the set of potential controls. Cases were eligible for selection as controls before their index date, and controls could be selected more than once [36].

See the Supplementary Material for the specification of the verification study cohort and the list of confounders that were considered.

Conditional logistic regression was used to estimate crude and confounder-adjusted odds ratios (ORs) with 95% confidence intervals (95% CI). We report ORs comparing current use of RVX to 1) current use of PPC (active comparator) and to 2) no exposure to RVX or PPC at index day. While the first should be used for a verification study, the second is needed to evaluate the decision made in the signal triage based on the Borda count ranks.

## 3 Results

In this section, we report the results of the case study. The results can be explored interactively on https://borda.bips.eu. The website also offers the opportunity to perform the same analysis with one’s own dataset.

### 3.1 Signal selection

Figure 2 shows Kendall’s tau correlations between ADR rankings of BCPNN, LGPS, LASSO and RF. It shows that these signal detection methods can yield quite different rankings. One example is the rank of N30.0 (acute cystitis); BCPNN, LGPS and RF rank this ICD-10-GM code under the top 2% of the strongest signals, whereas the LASSO ranks this ICD-10-GM code not even under the top 10%.

**Figure 2:**
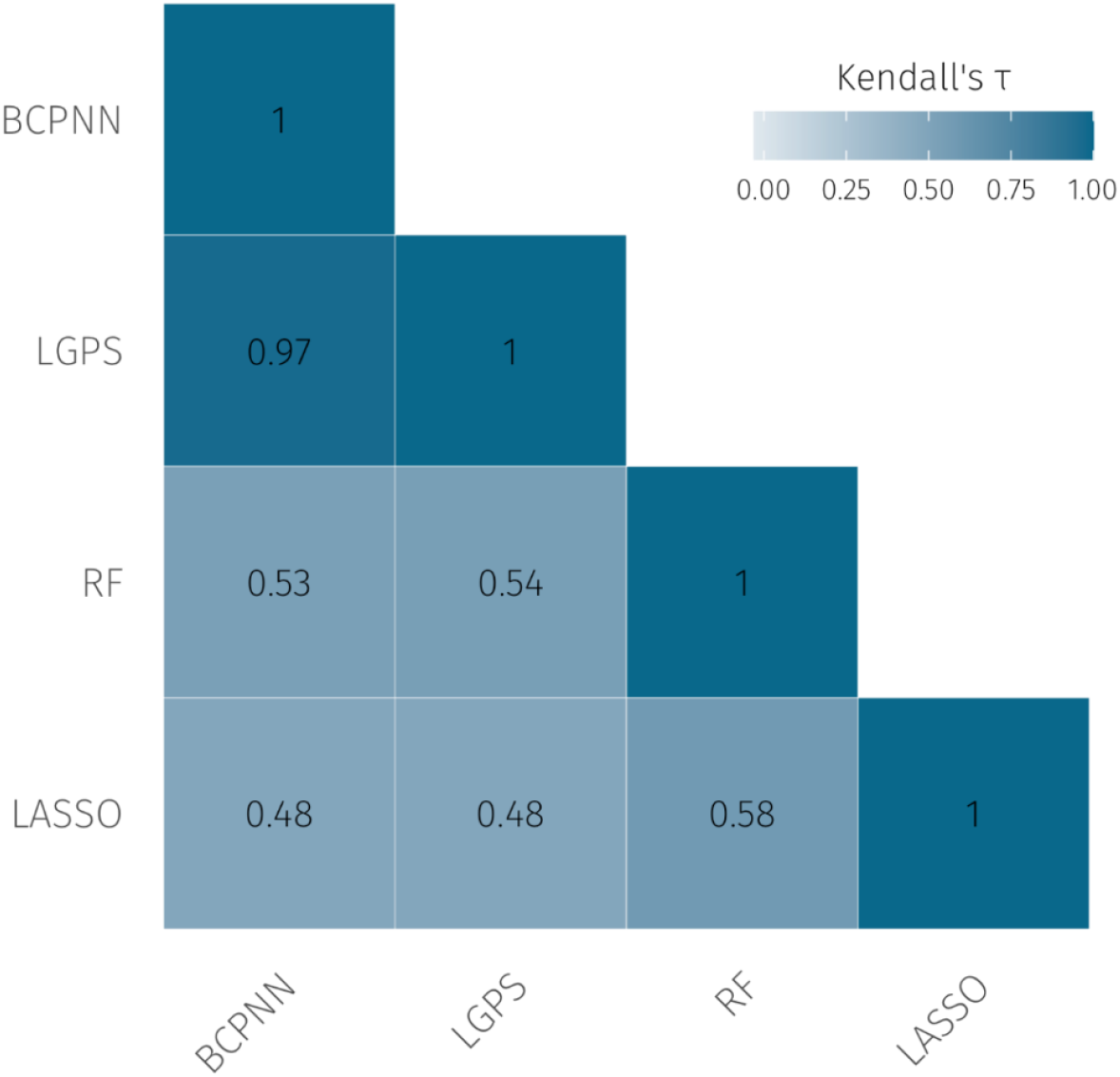
Kendall’s tau correlations between the rankings of the BCPNN, LGPS, RF and LASSO. The disproportionality methods, BCPNN and LGPS, lead to similar rankings. The LASSO and the RF, however, differ substantially from the others. We reduced the range of the legend for readability.

Table 2 shows the top 10 highest ranked ICD-10-GM codes according to the Borda ranking. Note that K92.2 (gastrointestinal hemorrhage, unspecified) is a known side effect of DOACs.

**Table 2:** Top ten ICD-10-GM codes for rivaroxaban according to the Borda ranking.

| Rank | ICD-10 code | Description |
| --- | --- | --- |
| 1 | D50.8 | Other iron deficiency anemias |
| 2 | J69.0 | Pneumonitis due to inhalation of food and vomit |
| 3 | K92.2 | Gastrointestinal hemorrhage, unspecified |
| 4 | K31.8 | Other specified diseases of stomach and duodenum |
| 5 | D50.0 | Iron deficiency anemia secondary to blood loss (chronic) |
| 6 | C20 | Malignant neoplasm of rectum |
| 7 | K25.0 | Acute gastric ulcer with hemorrhage |
| 8 | E86 | Volume depletion |
| 9 | I27.2 | Other secondary pulmonary hypertension |
| 10 | N17.9 | Acute kidney failure, unspecified |

### 3.2 Signal triage

On the basis of the ranking obtained during the signal detection phase, the committee selected 1) acute liver injury (ALI), 2) sepsis, and 3) acute cystitis (CYS). In addition, gastrointestinal bleeding (GB) and intracranial bleeding (ICB) were selected as positive controls.

This selection was made based on medical plausibility, whether the event was already known before, and with the help of plots as in Figure 3, in which the relative ranks for the ICD-codes related to the selected six health outcomes are shown. Each dot represents an individual ICD-code. A relative rank of “1” corresponds to the ADR to be on the bottom of the ranking (no signal). A relative rank of “0” means the item is on top of the ranking (strong signal). We prefer to use the relative rank for its readability. The ICD codes related to the positive controls GB and ICB are mainly on the top of the ranking as one would expect. The negative control, fracture of head and neck of femur, is clearly on the bottom of the ranking. The codes associated with ALI are close to the top of the ranking. Although many of the sepsis codes are at the bottom, the codes for streptococcal sepsis are noticeably clustered at the top of the ranking.

**Figure 3:**
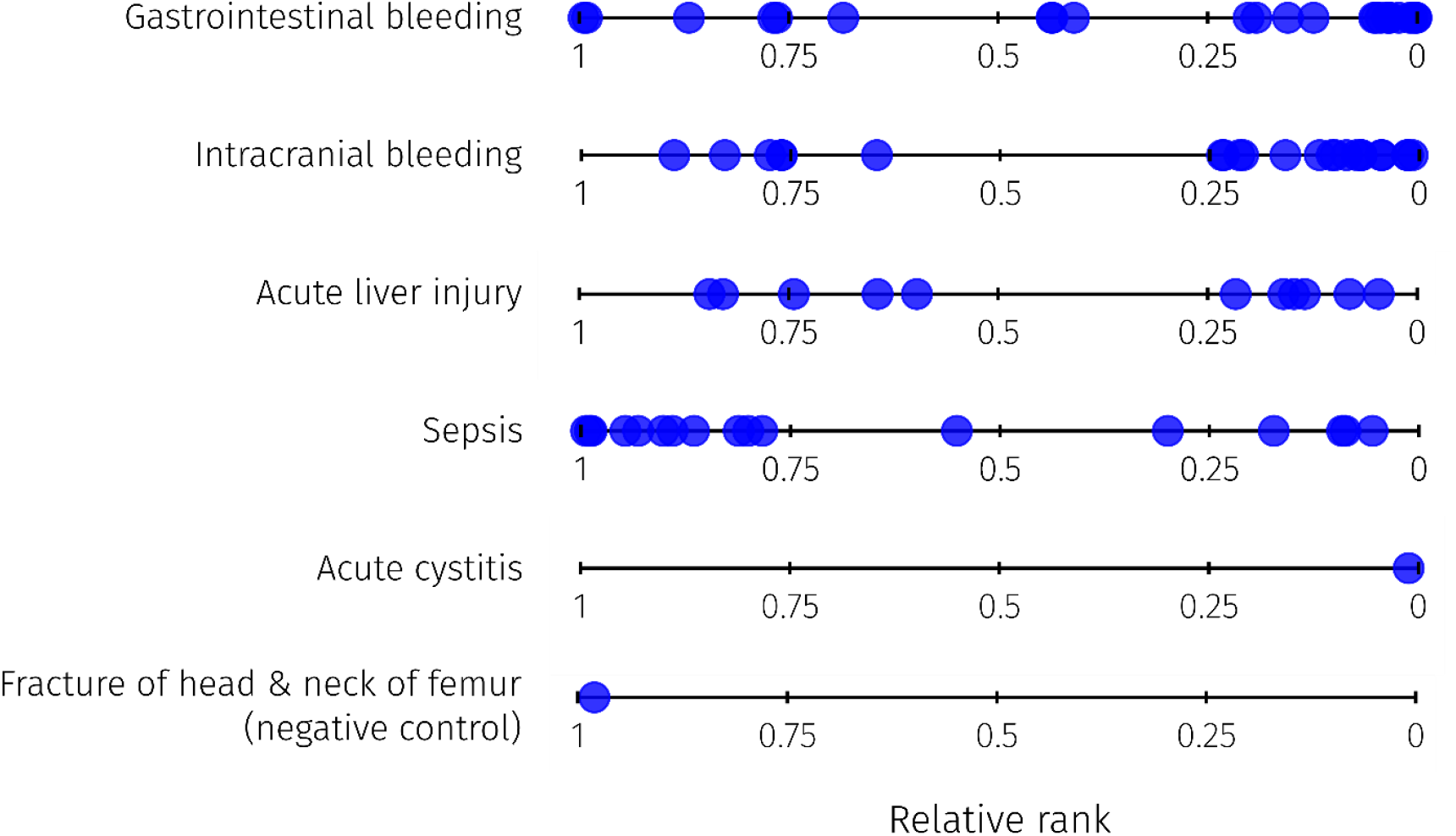
The relative ranks of events related to six health outcomes for rivaroxaban. A relative rank of 1 corresponds to the item being on the bottom of the list (no signal). A relative rank of 0 means the item is on top of the list (strong signal). Gastrointestinal and intracranial bleeding are known ADRs. Many of the ICD codes related to these outcomes are high up on the scale. The last, fracture of head and neck of femur (ICD-10: S720), is a negative control. The ICD code is, as expected, ranked close to the end. Acute liver injury and sepsis show both ICD codes that are ranked on the top as well as on the lower scale, whereas acute cystitis is ranked very high.

The ICD codes used to define the filtered signals (GB, ICB, ALI, CYS and sepsis) can be found in Table S1 in the Supplementary Material.

### 3.3 Verification study

The study cohort of the verification study was based on 97,400 new users of PPC and 71,917 users of RVX. The median age at cohort entry was 73 (inter-quartile-range (IQR): 65, 79) for RVX and 75 (IQR: 69, 81) for PPC new users. The proportion of women was 47% (RVX) and 48% (PPC). We observed 5053 cases of acute cystitis, 322 cases of acute liver injury, 3504 cases of sepsis, 6705 cases of gastrointestinal bleeding and 2974 cases of intracranial bleeding. A full description of the study cohort, including baseline characteristics, relevant medical history and dispensed medication at cohort entry, can be found in the Supplementary Material.

Table 3 shows that the risk of gastrointestinal bleeding and intracranial bleeding was increased in current users of RVX compared to no use (OR=2.45, 95% CI: (2.11; 2.84); OR=1.45, 95%CI: (1.18; 1.79)), but only the risk of gastrointestinal bleeding was increased compared to current use of PPC (OR=1.37, 95% CI: (1.27; 1.47); OR=0.74, 95% CI: (0.66; 0.83)). The risk of cystitis was slightly increased both in current users of RVX compared to no use and compared to current use of PPC (OR=1.12, 95% CI: (0.98; 1.29); OR=1.11, 95% CI: (1.02; 1.21)). The risk of acute liver injury was lower both in current users of RVX compared to no use and compared to current use of PPC (OR=0.76, 95% CI: (0.42; 1.37); OR=0.50, 95% CI: (0.35; 0.72)). The risk of sepsis was similar in current users of RVX compared to no use and compared to current use of PPC (OR=0.98, 95% CI: (0.82; 1.16); OR=1.10, 95% CI: (0.99; 1.23)).

**Table 3:**
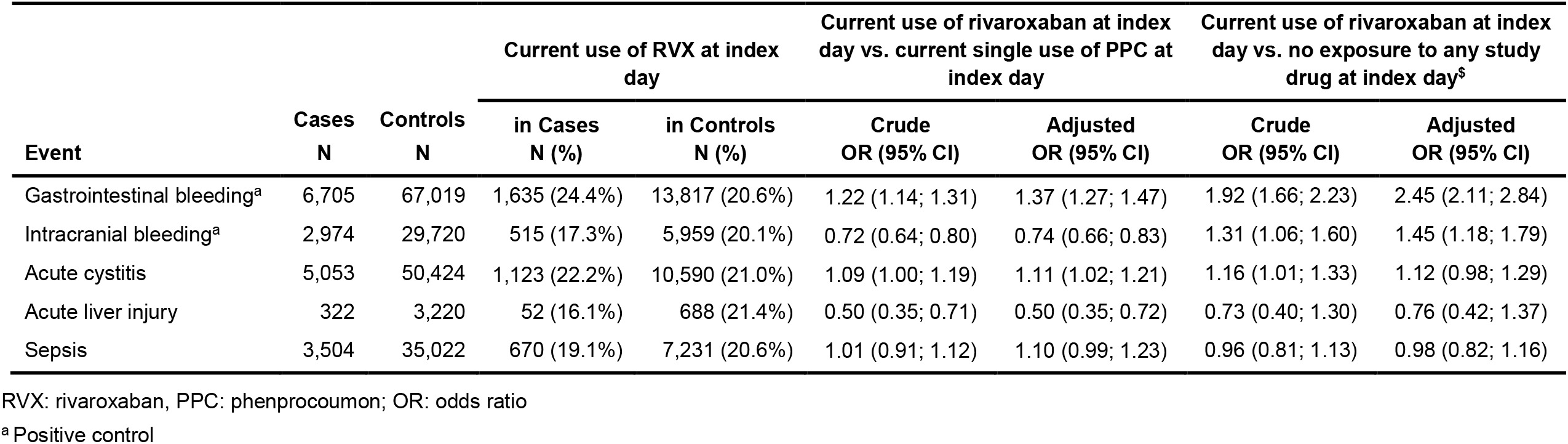
Results of the verification study: Crude and adjusted odds ratios and 95% confidence intervals

## 4 Discussion

In this paper we presented a novel three step approach for identifying and verifying potential adverse drug reactions in electronic health care data. In the common approach, a single signal detection method is applied and the resulting ranking of drug-ADR pairs is presented to a committee of medical experts. Here, we extend this process: First, we apply multiple signal detection methods simultaneously, where the obtained rankings are aggregated on the basis of the Borda count. Second, we offer the committee a graphical representation to define ADRs based on aggregated ICD codes. Third, we recommend to verify selected ADRs using a traditional pharmacoepidemiological approach. This can be of great help; the verification study might provide support for either the existence of the signal or its spuriousness.

We applied this novel approach in a case study for the DOAC rivaroxaban based on the health care claims dataset GePaRD. We applied four signal detection methods: two univariate disproportionality measures (BCPNN and LGPS), and two multivariate methods; on the one hand sparse regression (LASSO), and on the other hand a machine learning method (RF). We demonstrated that our proposed signal assessment was able to detect already known adverse events such as GB and ICB. It was also able to detect the rare event ALI. Although the verification study could not confirm that after choosing an appropriate study design and controlling for confounding current use of rivaroxaban was positively associated with ALI. The examples demonstrated that signal detection using health care claims data is feasible, but also that a verification is required to mitigate biases such as confounding, selection or measurement bias [37]. Here, we described signals of our case study that were either known before or signals that were not confirmed in the verification study. Additional results of our case study are described in [38], which investigates whether treatment of atrial fibrillation with direct oral anticoagulants affects the risk of epilepsy in comparison to treatment with phenprocoumon.

In general, the signal detection step is very fast and the inclusion of multiple detection methods may reduce overfitting. The verification study, however, is time consuming because it requires careful attention. Only a limited number of potential signals can hence be verified in a reasonable amount of time. Furthermore, the verification study should be enriched with several sensitivity analyses to investigate different sources of potential bias. It is also common practice to split the available data in a dataset for signal detection and for verification studies to avoid overoptimistic estimations of odds ratios and confidence intervals. In case little data is available, this could reduce the power to detect signals; important associations between drugs and ADRs might remain undetected. This might especially be the case when the drug of interest is prescribed infrequently, or the event occurs rarely.

Sandberg et al. [39] also presented a three-fold signal detection step, where first signals are estimated and then filtered by a list of criteria, followed by an in-depth signal assessment of the selected signals. Their approach focuses on subgroup disproportionalities and a catalog of criteria how to select signals for further assessment. Our approach focuses on a fast signal detection step which is based on various statistical methods and the combination of their results.

The advantage of aggregating rankings using Borda counts is that it does not depend on the interpretation of the scores produced by the individual methods. In addition, the rankings can be weighted, so that the results from various methods can differ in their contribution to the final ranking. This might be of interest, especially when one expects a certain method to perform exceptionally well in a particular instance. Adding more methods to the analysis is straightforward.

Working with electronic health care data has the downside that there tends to be a delay. As a consequence, ADRs of a newly admitted drug cannot be immediately detected. Here, spontaneous reporting systems certainly have an advantage over electronic health care data. However, post-market signal detection in claims data can complement spontaneous reports and is particularly useful a few years after first admission to the market, when spontaneous reports are less frequent and more electronic health care data are available.

## 5 Conclusions

We presented a novel approach to pharmacovigilance that allows the use of multiple signal detection methods simultaneously and offers the option to perform an additional verification study to the committee of medical experts. The case study shows that with this approach we are able to retrieve known ADRs and find other interesting signals. Performing a verification study, however, is time consuming but a severe, biologically plausible adverse event might demand immediate actions.

We developed an online tool available at https://borda.bips.eu that can aid in performing the analysis we presented here.

## Data Availability

All data produced in the present study are available upon reasonable request to the authors.

https://borda.bips.eu/

## Declarations

## Acknowledgments

The authors would like to thank all statutory health insurances which provided data for this study, namely AOK Bremen/Bremerhaven, DAK-Gesundheit, Die Techniker (TK), and hkk Krankenkasse. The authors would like to thank the data support team at the BIPS and Arne Godt for the implementation of the online tool.

## Data sharing

As we are not the owners of the data we are not legally entitled to grant access to the data of the German Pharmacoepidemiological Research Database. In accordance with German data protection regulations, access to the data is granted only to BIPS employees on the BIPS premises and in the context of approved research projects. Third parties may only access the data in cooperation with BIPS and after signing an agreement for guest researchers at BIPS.

## Code availability

The implementation of the methods that generated the signals presented in this paper is written in R and publicly available on github.com/bips-hb/pvm. We additionally used the R package ranger [1] for the random forest analysis.

## Authors’ contributions

Study concept and design: RF, LD (signal detection), MNW (signal detection), TS (verification study), IP Analysis and interpretation of data: RF, TS (verification study), MNW (signal detection), LD (signal detection), MS, TS (signal triage)

Drafting of manuscript: RF, TS, MNW, LD, IP

Critical revision of manuscript: RL, MNV

