## Supplementary Material for "A Discovery and Verification Approach for Pharmacovigilance using Electronic Health Care Data"

In this Supplementary Material, we will first describe the German Pharmacoepidemiological Research Database (GePaRD) which is the data source in this work. We give a complete description of the cohorts used in the signal detection and the verification phases in our case study. The ICD-10-GM codes used for the signals are presented in Table S1. The covariates used as confounders are given here as well.

### Data source (GePaRD)

We used GePaRD for the signal detection as well as for the verification phase of the case study. GePaRD is based on claims data from four statutory health insurance providers in Germany and currently includes information on approximately 25 million persons who have been insured with one of the participating providers since 2004 or later [1]. In addition to demographic data, GePaRD contains information on drug dispensations as well as outpatient (i.e., from general practitioners and specialists) and inpatient services and diagnoses. Per data year, there is information on approximately 20% of the living population and all geographical regions of Germany are represented. The suitability of GePaRD for pharmacoepidemiological research has been demonstrated by methodological as well as various types of pharmacoepidemiological studies investigating the safety of drugs [1].

In Germany, transferring health insurance data for scientific research without asking for informed consent is regulated by the Code of Social Law. All involved health insurance providers as well as the German Federal Office for Social Security and the Senator for Health, Women and Consumer Protection in Bremen as their responsible authorities approved this study according to the legal conditions. According to the Ethics Committee of the University of Bremen studies based on GePaRD are exempt from institutional review board review.

For more information, please visit the webpage: https://www.bips-institut.de/en/research/research-infrastructures/gepard.html

### Cohort signal detection study

The study cohort for the signal detection phase of our case study contained all persons who

1. had been insured with the health insurance Techniker Krankenkasse (TK) for at least 12 consecutive months between January 2015 and December 2016;

2. had at least one dispensation of rivaroxaban between 2015-04-02 and 2016-10-02, i.e., at least 90 days after cohort entry and before cohort exit, respectively, and
3. had no DOAC use within 12 months preceding cohort entry.

Persons were excluded from the study cohort if they switched to another DOAC. We use 4-digit ICD-10-GM codes for detecting potential signals.

### A self-controlled case series design

A self-controlled case series design [2] was used for the LASSO and the RF in the signal detection phase of our case study. See Figure S1 for a graphical representation.

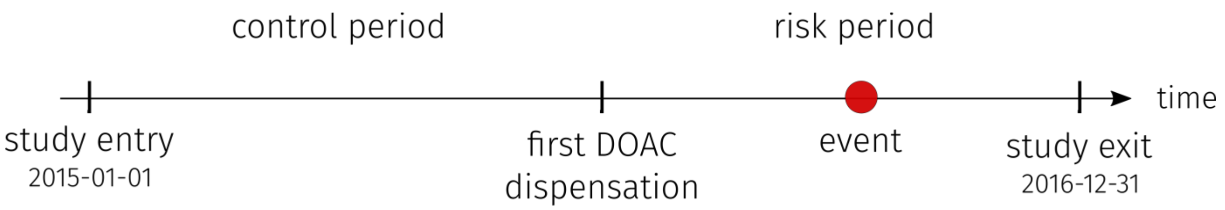


**Figure S1:** The self-controlled case series design. Shown is a representation for a single patient from the start of 2015 till the end of 2016, respectively, the study entry and study exit. The control period is the time between entry and the first DOAC dispensation; the risk period is between the first dispensation and study exit. Here, the patient suffered the adverse event during the risk period.

### The ICD-10-GM codes for health outcomes

The ICD-10-GM codes used in the verification phase in our case study for the health outcomes can be found in Table S1.

**Table S1:** ICD-10-GM codes used for health outcome definition

| Health outcome | ICD-10-GM |
|---|---|
| Gastrointestinal bleeding | I98.3, K22.6, K22.8, K22.80, K22.81, K22.88, K25.0, K25.2, K25.4, K25.6, K26.0, K26.2, K26.4, K26.6, K27.0, K27.2, K27.4, K27.6, K28.0, K28.2,K 28.4, K28.6, K29.0, K31.8, K55.2, K55.3, K55.8, K57.0, K57.1, K57.2, K57.3, ,K57.4 , K57.5, K57.8, K57.9, K62.5, K66.1, K92.0, K92.1, K92.2 |
| Intracranial bleeding | I60.0, I60.1, I60.2, I603, I60.4, I60.5, I60.6, I60.7, I60.8, I60.9, I61.0, I61.1, I61.2, I61.3, I61.4, I61.5, I61.6, I61.8, I61.9, I62.0, I62.00, I62.01, I62.02, I62.09, I62.1, I62.9, S06.3, S06.4, S06.5, S06.6 |
| Acute liver injury | K71.0, K71.1, K71.2, K71.6, K71.7, K71.8, K71.9, K72.0, K72.9, K75.2, K75.3, K75.8, K75.9, K76.2 |
| Acute cystitis | N30.0 |
| Sepsis | A02.1, A20.7, A22.7, A26.7, A32.7, A39.2, A39.3, A39.4, A40.0, A40.1, A40.2, A40.3, A40.8, A40.9, A41.0, A41.1, A41.2, A41.3, A41.4, A41.5, A41.5, A41.8, A41.9, A42.7, B37.7 |

### Cohort verification study

The study cohort for the verification phase of our case study included all persons from January 2011 to December 2017 who 1) had at least one dispensation of RVX or PPC during the study period, 2) had been continuously insured for at least 12 months before cohort entry, 3) had no dispensing of RVX or PPC in the 12 months before cohort entry ("new user"), 4) had at least one diagnosis of atrial fibrillation within 12 months before cohort entry, and 5) had no diagnosis of cancer any time before cohort entry and were not pregnant at cohort entry. Exposure at index date was defined as a supply overlapping the index date, whereas supply was estimated as the number of defined daily doses of the last dispensing before the index day.

Only hospital diagnoses were considered for the events acute liver injury, intracranial bleeding, gastrointestinal bleeding and sepsis. The index date was set to the respective admission date and patients with a respective diagnosis in the 180 days before cohort entry were excluded. For cystitis, both outpatient and hospital diagnoses were taken into account. The index date was set to the date of diagnosis or the admission date and all patients with a respective diagnosis in the 365 days before cohort entry were excluded.

The study cohort of the verification study was based on 97,400 new users of PPC and 71,917 users of RVX. The median age at cohort entry was 73 (IQR: 65, 79) for rivaroxaban and 75 (IQR: 69, 81) for PPC new users. The proportion of women was 47 % (RVX) and 48 % (PPC). We observed 6705 cases of gastrointestinal bleeding, 2974 cases of intracranial bleeding, 322 cases of acute liver injury, 5053 cases of acute cystitis and 3504 cases of sepsis. Baseline characteristics including relevant medical history and dispensed medication at cohort entry are presented in Table S2.

### Confounders included in the verification study

Age, risk scores for bleeding (HAS-BLED) and stroke ($CHA_2DS_2$-VASC), risk factors for the respective events, other comorbidities, lifestyle factors, and comedication were assessed at baseline (with appropriate lookback periods) as potential confounders or effect modifiers. Comedication was additionally assessed on index date. The quintiles of the German Index of Socioeconomic Deprivation of the place of living were used as proxy for the socio-economic status and access to health care.

**Table S2:** Characteristics of cases and matched controls at cohort entry

| Characteristics | Cases n (%) | Controls n (%) |
|---|---|---|
| **Gastrointestinal bleeding** | 6,705 | 67,019 |
| Females | 3,531 (53%) | 35,296 (53%) |
| Age: *median (IQR)* | 80.0 (74.0, 85.0) | 80.0 (74.0, 85.0) |
| ***Medical history*** | | |
| Acute renal failure | 54 (0.8%) | 189 (0.3%) |
| Heart failure | 4,243 (63%) | 34,330 (51%) |
| Hypertension | 6,564 (98%) | 64,057 (96%) |
| Moderate/severe chronic kidney disease | 1,740 (26%) | 10,862 (16%) |
| Liver disease | 2,343 (35%) | 18,931 (28%) |
| ***Medication use*** | | |
| Antithrombotics | 1,534 (23%) | 11,275 (17%) |
| CYP-/P-glycoprotein inducers[1] | 30 (0.4%) | 233 (0.3%) |
| **Intracranial bleeding** | 2,974 | 29,720 |
| Females | 1,497 (50%) | 14,970 (50%) |
| Age: *median (IQR)* | 80.0 (74.0, 85.0) | 80.0 (74.0, 85.0) |
| Alcohol and drug abuse | 186 (6.3%) | 1,235 (4.2%) |
| ***Medical history*** | | |
| Brain disease | 58 (2.0%) | 244 (0.8%) |
| Ischemic stroke | 628 (21%) | 4,699 (16%) |
| Transient ischemic attack | 168 (5.6%) | 1,548 (5.2%) |
| Moderate/severe chronic kidney disease | 584 (20%) | 4,764 (16%) |
| ***Medication use*** | | |
| Antithrombotics | 553 (19%) | 5,099 (17%) |
| CYP-/P-glycoprotein inducers[1] | 14 (0.5%) | 125 (0.4%) |
| Systemic corticosteroids | 605 (20%) | 5,479 (18%) |
| Pethidine, Tramadol | 320 (11%) | 3,299 (11%) |
| **Acute liver injury** | 322 | 3,220 |
| Females | 184 (57%) | 1,840 (57%) |
| Age: *median (IQR)* | 75.0 (69.0, 80.0) | 75.0 (69.0, 80.0) |
| Alcohol and drug abuse | 31 (9.6%) | 149 (4.6%) |
| ***Medical history*** | | |
| Cardiovascular disease | 203 (63%) | 1,957 (61%) |
| Diabetes mellitus | 146 (45%) | 1,029 (32%) |
| Chronic hepatitis | 4 (1.2%) | 18 (0.6%) |
| Other liver disease | 139 (43%) | 919 (29%) |
| ***Medication use*** | | |
| Antibiotics[1] | 9 (2.8%) | 46 (1.4%) |
| CYP-/P-glycoprotein inducers[1] | 3 (0.9%) | 17 (0.5%) |
| NSAIDs | 256 (80%) | 2,619 (81%) |
| **Cystitis** | 29,698 | 296,532 |
| Females | 18,885 (64%) | 188,535 (64%) |
| Age*: median (IQR)* | 77.0 (71.0, 83.0) | 77.0 (71.0, 83.0) |
| Alcohol and drug abuse | 1,405 (4.7%) | 12,323 (4.2%) |
| ***Medical history*** | | |

| Characteristics | Cases n (%) | Controls n (%) |
|---|---|---|
| Genital abnormalities | 4,756 (16%) | 38,456 (13%) |
| Other disorders related to cystitis | 7,445 (25%) | 68,339 (23%) |
| Prostatic disease | 6,825 (23%) | 59,606 (20%) |
| Urinary disorders | 8,986 (30%) | 59,716 (20%) |
| **Sepsis** | 3,504 | 35,022 |
| Females | 1,529 (44%) | 15,286 (44%) |
| Age: *median (IQR)* | 79.0 (73.0, 84.0) | 79.0 (73.0, 84.0) |
| Alcohol and drug abuse | 264 (7.5%) | 1,506 (4.3%) |
| ***Medical history*** | | |
| Anemia | 1,628 (46%) | 10,577 (30%) |
| Heart failure | 2,368 (68%) | 17,228 (49%) |
| Hypertension | 3,445 (98%) | 33,370 (95%) |
| Malnutrition | 117 (3.3%) | 376 (1.1%) |
| Moderate/severe chronic kidney disease | 1,146 (33%) | 5,368 (15%) |
| ***Medication use*** | | |
| Antiinfective drugs[a] | 261 (7.4%)2 | 532 (1.5%)2 |
| CYP-/P-glycoprotein inducers[a] | 23 (0.7%)2 | 123 (0.4%)2 |
| Immunosuppressants | 203 (5.8%) | 777 (2.2%) |

Values are numbers (percentages) unless stated otherwise
[a] Supply was overlapping the index
